# Population prevalence, penetrance, and mortality for genetically confirmed MODY

**DOI:** 10.1101/2025.06.30.25330354

**Authors:** Luke N Sharp, Kevin Colclough, Jacques Murray Leech, Stuart J Cannon, Thomas W Laver, Andrew T Hattersley, Michael N Weedon, Kashyap A Patel

**Affiliations:** Department of Clinical and Biomedical Sciences, Faculty of Health and Life Sciences, University of Exeter, UK; Exeter Genomic Laboratory, Royal Devon University Healthcare NHS Foundation Trust, Exeter, UK

## Abstract

**Context:** Diagnosing Maturity-Onset Diabetes of the Young (MODY) is clinically important for treatment and prognosis. However, phenotype-based studies of MODY are prone to ascertainment bias, limiting accurate estimates of its population prevalence and phenotypic spectrum.

**Objective:** To apply a genotype-first approach to determine the population prevalence, penetrance, and all-cause mortality associated with MODY.

**Methods:** We analysed exome sequencing and clinical data from 454,275 UK Biobank participants to identify pathogenic variants in 10 established MODY genes. We assessed variant prevalence, age-dependent diabetes penetrance, and all-cause mortality by genetic aetiology over a mean follow-up of 13.4 years.

**Results:** Pathogenic MODY variants were present in 1 in 1,052 individuals and accounted for 1.48% of diabetes cases diagnosed before age 40. *GCK* variants were the most frequent (1 in 2,787), demonstrating high penetrance (mean HbA1c 8.8 mmol/mol higher; 94.5% with prediabetes or diabetes) but no significant association with all-cause mortality (*P*=0.09). Variants in other MODY genes showed lower penetrance, with 12% of carriers developing diabetes by age 40 and 31.6% by age 60 and showed no increase in all-cause mortality (*P*=0.89). Penetrance varied by genetic aetiology, with *HNF1A* showing the highest penetrance and *PDX1*, *NEUROD1*, and *RFX6* the lowest. Parental history of diabetes and polygenic risk for type 2 diabetes were important modifiers of penetrance (Hazard ratios 2.54 and 1.52 respectively, *P*<3.9×10^−3^).

**Conclusions:** This large-scale genotype-first study provides novel insights into MODY in the population. These findings have broad implications for genetic counselling, personalised treatment strategies, and healthcare resource allocation.

## Introduction

Our current understanding of Maturity-Onset Diabetes of the Young (MODY) particularly its prevalence, penetrance, and impact on mortality has largely been shaped by studies using a phenotype-first approach. MODY is an autosomal dominant form of monogenic diabetes caused by pathogenic variants in at least 11 genes (1,2). Accurate diagnosis is clinically important, as it directly informs treatment decisions and prognosis (1,3,4). Most studies on MODY have focused on young-onset diabetes that lack biomarkers for type 1 diabetes or familial diabetes identified from the routine clinical practice which are later confirmed to have genetic diagnosis of MODY (5–9). These studies are referred as phenotype-first studies. Due to this reliance on the phenotype selection, studies of monogenic disorders have shown to suffer from ascertainment bias (5–11). These may lead to overestimation of age-dependant penetrance by focusing on individuals who develop diabetes at a young age (10). Similarly, due to homogenous selection criteria, these studies are potentially less powerful to assess the difference in phenotype due to underlying genetic aetiology. The ascertainment bias of phenotype first studies may also potentially explain the excess of female cases in genetically confirmed MODY (4,8,12).

In contrast, genotype-first studies, which identify individuals based on genetic variants irrespective of phenotype, offer a powerful way to overcome the limitations of phenotype-first studies (11,13). The lack of selection of the phenotype means they are well-suited to assessing the full clinical spectrum, prevalence and outcomes such as all-cause mortality, which remains unknown for most MODY subtypes. This is particularly relevant for *GCK*-MODY, which has a unique phenotype of lifelong mild fasting hyperglycaemia, the effect of which on mortality has not been established. The previous genotype-first studies in MODY have been limited by restricted gene coverage (10), lack of mortality data (10,14,15), lack of age-dependent diabetes risk (14,15), or reliance on case-control designs (15) and lack the assessment of factors affecting penetrance of diabetes (10,14,15) constraining their ability to provide comprehensive, population-level insights.

In this study, we leverage a genotype-first approach in a large population cohort from the UK to provide novel insights into MODY in the population. We aim to investigate the prevalence, age-dependent penetrance of diabetes, factor affecting penetrance, and all-cause mortality associated with MODY in a cohort of 454,275 individuals from the UK population.

## Research Design and Methods

### Study Population

To assess the prevalence, penetrance, and all-cause mortality in the population, we used data from the UK Biobank, a large population-based study in the United Kingdom with participants the aged between 40-70yrs at recruitment (16). These participants provided detailed phenotypic information through electronic health records, self-reported questionnaires, and structured interviews. They also measured biomarkers, including key diabetes-related markers such as HbA1c and glucose from the blood. Importantly, all participants also had comprehensive genetic data available. For this study, we analysed whole exome sequencing from the 450,000 data release from the UK Biobank(16,17). Supplemental Table S1 provides the characteristics of the individuals included in the current study. Ethics approval for the UK Biobank study was obtained from the North West Centre for Research Ethics Committee (11/NW/0382). Written informed consent was obtained from all participants.

### Diabetes and all-cause mortality

We defined individuals with diabetes if they self-reported to have diabetes, were on diabetes medication or had an HbA1c level > 48 mmol/mol (6.5%) at the time of recruitment into the study. We incorporated self-reported age of diabetes diagnosis and the first occurrence data into our analysis. We defined prediabetes based on the American Diabetes Association (ADA) criteria, using a fasting glucose level ≥ 5.6 mmol/L or an HbA1c level ≥ 39 mmol/mol (18).

We obtained all-cause mortality data up to November 2022 through the National Death Registries Linkage, which the UK Biobank carried out centrally. Detailed information is available at: https://biobank.ndph.ox.ac.uk/ukb/refer.cgi?id=115559.

### Genetic Data

We utilised the 454,275 whole exome sequencing release from the UK Biobank. *Szustakowski et al.* described the detailed methodology for this sequencing, which is available at https://biobank.ctsu.ox.ac.uk/showcase/label.cgi?id=170 (17). Briefly, the IDT xGen Exome Research Panel v1.0 was used which covered the exons of 19,396 genes. Samples were sequenced using the Illumina NovaSeq 6000 platform with mean coverage exceeding 20x at 95.2% of sites. Variants were called to the GRCh38 genome build reference sequence using the OQFE protocol (17).

We generated the type 1 diabetes genetic risk score (T1DGRS) and type 2 diabetes polygenic risk score (T2DPRS) using TOPMed imputed data genotyping array data released by the UK Biobank. Detailed information on this is available at https://biobank.ndph.ox.ac.uk/ukb/field.cgi?id=21007 and described by Arni *et al.* (19,20).

We used a genome-wide T2DPRS developed by Huerta-Chagoya *et al.*, which was derived from individuals excluding UK Biobank participants (21). To capture type 1 diabetes polygenic risk, we employed the T1DGRS2 developed by Sharp *et al.* (19,21,22).

### Classification of MODY variants

We analysed variants in the coding regions of 10 genes known to cause MODY: *ABCC8*, *GCK*, *HNF1A*, *HNF1B*, *HNF4A*, *INS*, *NEUROD1*, *PDX1*, *RFX6*, *KCNJ11* (2). We excluded *CEL*-MODY from this analysis as it is not possible to call the single base deletions that cause *CEL*-MODY from whole exome sequencing data as they occur in the gene’s variable number tandem repeat region (23,24). We annotated these variants using Ensembl’s Variant Effect Predictor (25). We classified as pathogenic or likely pathogenic according to ACMG/AMP guidelines (26,27). To minimise the inclusion of false-positive variants, we only included protein-truncating variants (PTVs) in the study if they were classified as high confidence by LOFTEE (28). To further limit the inclusion of very low-penetrance variants, we only included pathogenic missense variants if they were ultra-rare in the population (maximum allele count of 2 in gnomAD v2.1.1, MAF <1.4×10^−5^). We manually reviewed sequence read data for all pathogenic protein-truncating variants using the Integrative Genomics Viewer (IGV) to exclude false-positive variants (29). The variants included in the analysis are listed in Supplemental Table S2. We also included individuals with *HNF1B*-MODY caused by deletions in the 17q12 region, as nearly half of the *HNF1B*-MODY cases are due to this deletion (30). We used genotyping array data for this analysis, as described in previous work by Cannon *et al.* (30).

### Statistical Analysis

We statistically analysed proportions/counts between categories and groups using the Fisher’s exact test. To calculate 95% confidence intervals (CIs) for proportions, we applied the exact binomial confidence interval Clopper–Pearson method. We used Kaplan-Meier survival analysis to estimate age-dependent penetrance of diabetes and all-cause mortality. To compare diabetes penetrance and all-cause mortality between groups, we performed the log-rank test. We computed hazard ratios for diabetes and all-cause mortality using Cox proportional hazard models, both with and without adjustment for covariates. We assessed the impact of age on HbA1c levels through linear regression. Welch two sample T-tests were used to compare HbA1c and blood glucose levels between *GCK* heterozygotes and non-carriers. To compare the penetrance of mild hyperglycaemia in *GCK* heterozygotes between cohorts and to test the difference in prevalence between males and females, we applied Fisher’s exact tests. We evaluated heterogeneity in prevalence by ancestry using Cochran’s Q test using STATA package META. We conducted all analyses using STATA 18.0 and R version 4.2.2 with RStudio.

## Results

### At least 1 in 1,052 individuals in the UK Biobank carries a pathogenic MODY variant, with *GCK* variants being the most common seen in 1 in 2,787 individuals

Our analysis of variants in 10 MODY genes across 454,275 UK Biobank participants identified 432 individuals carrying pathogenic MODY variants. This represents a prevalence of 0.095% (95% CI 0.086-0.1, 1:1052) (Figure 1A). Pathogenic variants in *GCK* were most prevalent, comprising 37.7% (163/432) of identified carriers, followed by pathogenic variants in *RFX6* at 22% (95/432) (Figure 1B). This population prevalence of *GCK*-MODY was 1:2787 (0.036%, 95% CI 0.031-0.042%). Among individuals with diabetes at recruitment, 0.63% carried pathogenic MODY variants (95% CI 0.54-0.73%), 181/28,795, 1 in 159). The prevalence of MODY was 1 in 68 for those diagnosed under age 40yrs (1.48%, 95% CI 1.06-2.01%, 40/2,706), and 1 in 141 to those diagnosed before age 60yrs (0.71%, 95% CI 0.6-0.84%, 136/19,187). Unlike clinically selected cohorts, male and female individuals with diabetes showed similar prevalence rates (0.095% vs 0.095%, *P*=1). The prevalence of pathogenic MODY variants remained similar across European (0.096%), South Asian (0.12%), and other (0.091%) ancestries, though individuals of African ancestry showed significantly lower prevalence (0.014%) (*P heterogeneity* 2.1×10^−7^) (Supplemental Table S3).

**Figure 1:**
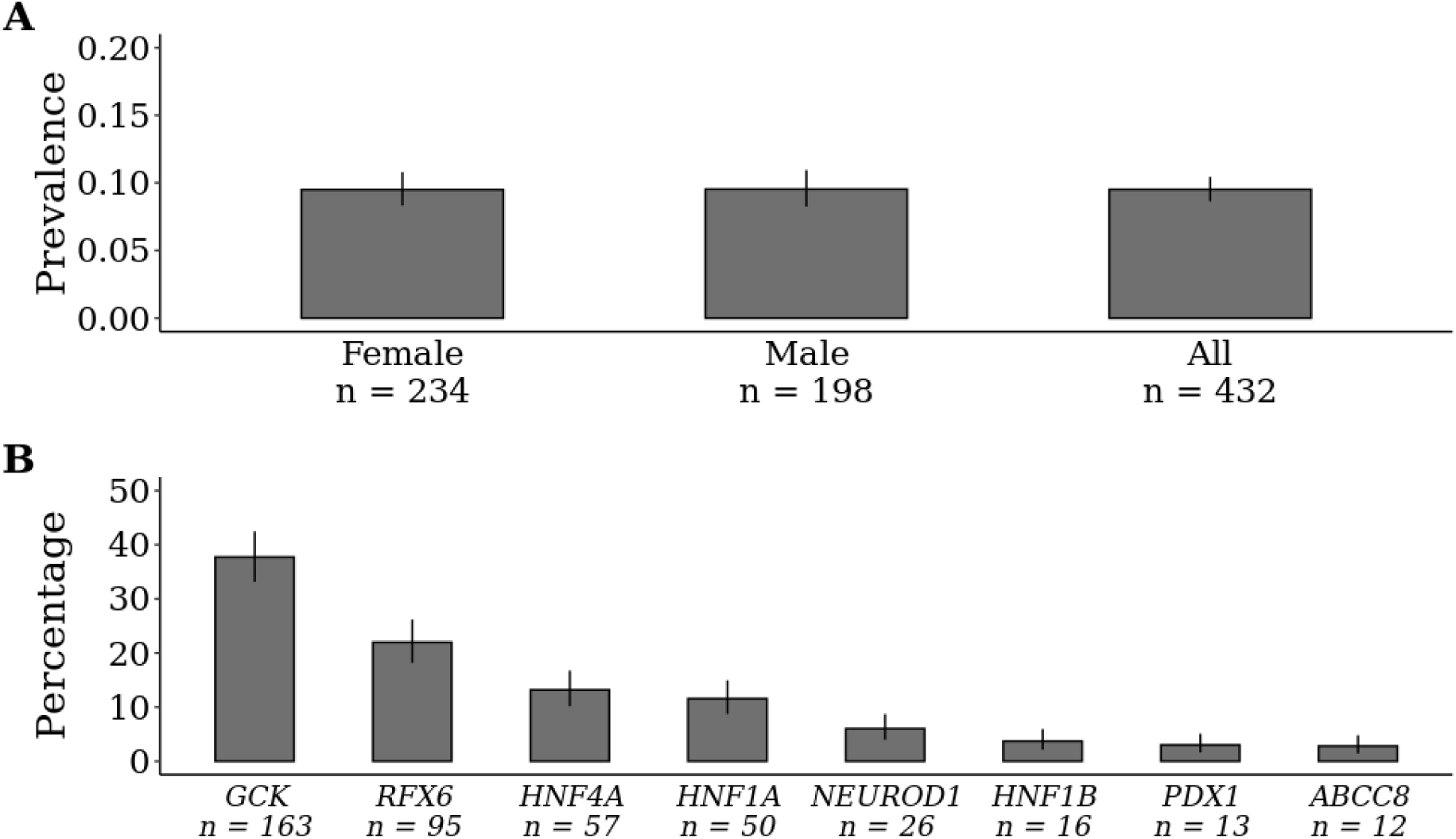
**Prevalence of pathogenic variants in 10 MODY genes in a population cohort.** A) The graph shows the prevalence (%) of pathogenic variants associated with 10 MODY genes across 454,275 individuals from the UK biobank (n=432 carriers), with data stratified by sex (female, 234/246,437 and male 198/207,838). B) The distribution of pathogenic variants among the 432 carriers from the UK Biobank, categorised by genetic aetiology. Error bars indicate 95% confidence intervals.

### Pathogenic *GCK* variants show high penetrance in a population cohort

Pathogenic variants in *GCK* have been shown to cause mild lifelong fasting hyperglycaemia in a clinically ascertained cases (31). We found that carriers of pathogenic *GCK* variants showed on average an 8.7 mmol/mol higher HbA1c levels compared with non-carriers (47.1+/-SD4.9 vs 38.4+/-6.4 mmol/mol, *P*=4.02×10^−51^) (Figure 2A) (Supplemental Table S4). Similarly, their glucose after at least 5 hours of fasting was 1.28 mmol/L higher compared with non-carriers (6.34+/-0.97 vs 5.06 +/-1.04 mmol/L, *P*=3.41×10^−10^) (Supplemental Table S4). The *GCK* variant carriers showed an age-dependent increase in HbA1c (beta 0.17, *P*=7.17×10^−4^), which was similar to non-*GCK* carriers (*P* interaction 0.76). The rate of prediabetes in pathogenic *GCK* variant carriers was 94.5% (95% CI 89.8-97.4%) compared with 34.3% (95% CI 34.2-34.4%) in non-carriers (*P*=1.21×10^−59^) (Figure 2B).

**Figure 2:**
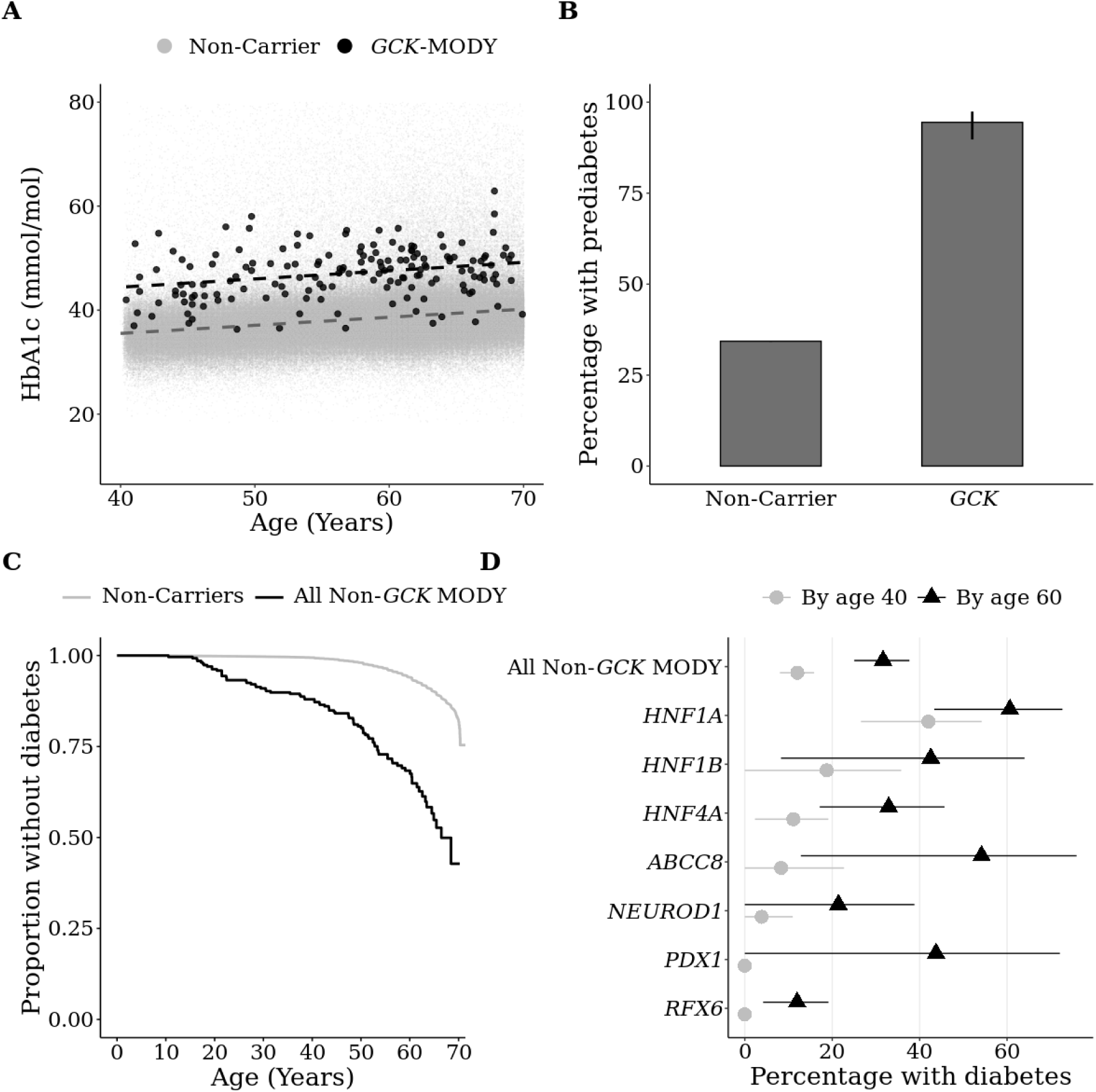
**Penetrance of pathogenic MODY variants in the population cohort** A) Scatter plot illustrating the relationship between HbA1c (mmol/mol) and age at recruitment, stratified by pathogenic *GCK* variant carrier status. Dashed lines represent linear trends for each group. B) Bar chart displaying the percentage of individuals with prediabetes, with error bars indicating 95% confidence intervals. C) Kaplan-Meier analysis of diabetes onset in UK Biobank participants, stratified by pathogenic MODY variant carrier status, excluding individuals carrying pathogenic variants in *GCK*, (Non-*GCK*-MODY n=163). D) The proportion of pathogenic MODY variant carriers developing diabetes by age 40 (grey) and age 60 (black), with 95% confidence intervals.

### Non-*GCK* pathogenic MODY variants demonstrate variable age-dependent diabetes penetrance across genetic aetiologies

We next assessed the age-dependent risk of diabetes in carriers of pathogenic variants in non-*GCK* MODY genes within our population cohort. By age 40yrs, 12% (95% CI 8-15.9%) of non-*GCK* pathogenic MODY variant carriers had developed diabetes, increasing to 31.6% (95% CI 25-37.7) by age 60yrs (Figure 2C). These variants together increased diabetes risk by nearly seven-fold (HR 6.86, 95% CI 5.55-8.48, *P*=4.19×10^−71^). However, the diabetes penetrance varied substantially across genetic aetiologies (Figure 2D). *HNF1A* pathogenic variants emerged as the most penetrant cause of diabetes, with 42% (95% CI 26.6-54.2) of carriers developing diabetes by age 40yrs, rising to 60.6% (95% CI 43.3-72.7) by 60yrs. In contrast, *RFX6*, *PDX1* and *NEUROD1* pathogenic variants showed the lowest penetrance. No *RFX6* or *PDX1* carriers had diabetes by age 40yrs while only 3.8% of *NEUROD1* carriers had diabetes. By 60yrs 12% of *RFX6* carriers, 43.8% of *PDX1* carriers and 21.4% of *NEUROD1* carriers had diabetes (Figure 2B) (Supplemental Table S5).

### Type 2 diabetes polygenic score and parental diabetes status affect the penetrance of pathogenic MODY variants

Our genotype-first approach allowed us to assess the true impact of environmental and polygenic factors on age-dependent diabetes penetrance. We found that sex, BMI and type 1 diabetes genetic risk had no effect on diabetes penetrance in carriers of pathogenic non-*GCK* MODY variants (Table 1). However, parental history of diabetes increased the diabetes risk ∼2-fold (hazard ratio 2.54, 95% CI 1.56-4.14), and each standard deviation increase in type 2 diabetes polygenic risk score increased the risk by 1.52-fold (95% CI 1.14-2.03, *P*=3.9×10^−3^) (Table 1). These results were directionally consistent across genetic aetiologies but were limited by small numbers in each group (Supplemental Table S6).

**Table 1:**
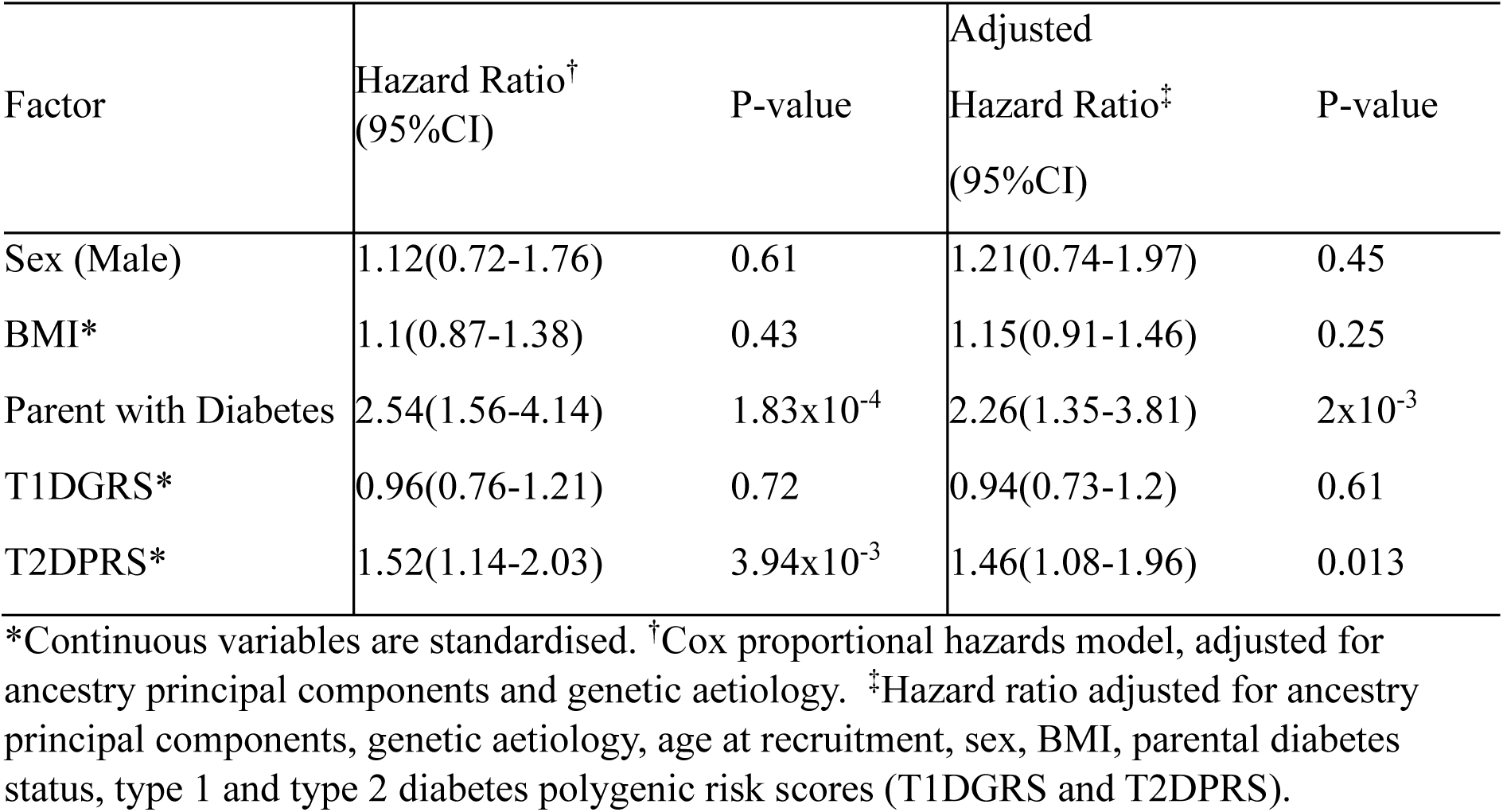
Factors influencing diabetes penetrance in pathogenic non-*GCK* MODY pathogenic variant carriers.

### Pathogenic MODY variants carriers do not show excess all-cause mortality

Finally, we assessed whether the carriers of pathogenic variants were at higher risk of all-cause mortality. Our Kaplan-Meier analysis did not demonstrate excess all-cause mortality for non-*GCK* MODY carriers compared with non-carriers over mean follow up of 13.4 years at mean age of 70.4 years (log-rank *P* 0.89) (Figure 3A). Similarly, despite higher HbA1c levels, pathogenic *GCK* variant carriers did not show excess all-cause mortality (log-rank *P* 0.09) (Figure 3B). The all-cause mortality for *GCK* pathogenic variant carriers remained similar, regardless of whether they had diabetes diagnosed by a doctor, were being administered statins, or had HbA1c levels in the highest or lowest tertiles in the carriers (All log-rank *P* > 0.4) (Supplemental Figure 1). The results were also consistent across all MODY subtypes, both individually and combined (*P* > 0.05) and after adjusting for sex, smoking status, diabetes status, age at recruitment and HbA1c (Figure 3C) (Supplemental Table S7).

**Figure 3:**
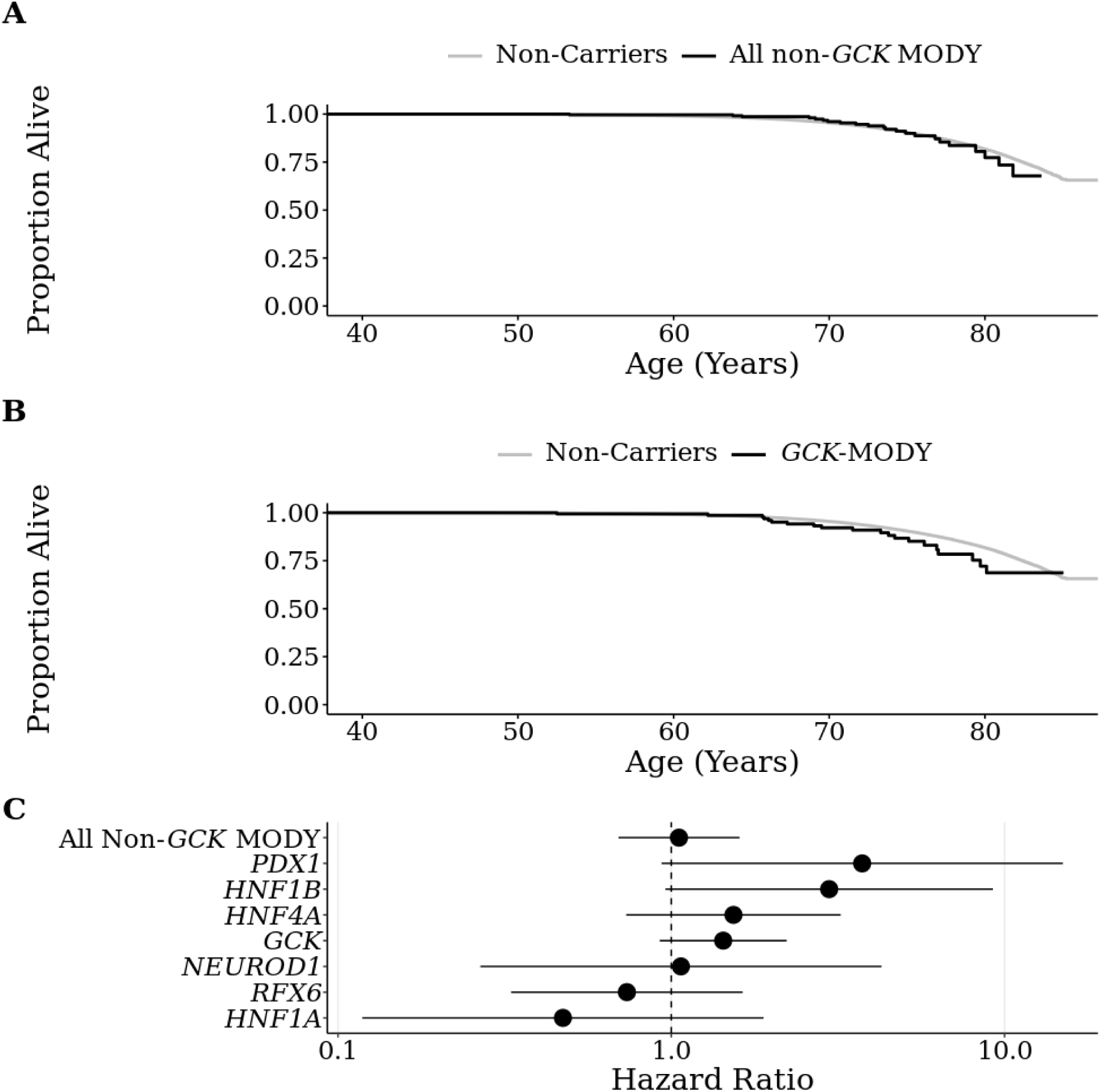
**All-cause mortality in pathogenic MODY variant carriers in a population cohort** Kaplan-Meier analysis of all-cause mortality in 454,275 participants from the UK biobank split by A) pathogenic non-*GCK* variant carriers (n=269) and non-carriers and B) pathogenic *GCK* variant carriers (n=163) and non-carriers. C) A forest plot showing the hazard ratios for mortality split by genetic aetiology adjusted for genetic ancestry principal components. *ABCC8* has not been included due no mortality occurring in *ABCC8* variant carriers.

## Discussion

In this large population study using a genotype-first approach, we show that MODY is prevalent in the population (1:1052) and accounts for 1.48% of diabetes under 40 years. The penetrance is much lower than previously estimated and varies substantially by genetic aetiology, family history and polygenic type 2 diabetes background. We also show that MODY does not cause an excess all-cause mortality, even in individuals with *GCK*-MODY who have life-long excess glucose burden.

Our genotype-first approach with a large sample size provides novel insights over phenotype-first studies. We report prevalence of *GCK*-MODY of 1:2787 (0.036%). This is lower than the currently widely adopted estimate of ∼1:1000 by Chakera *et al*. (32). However, this estimate is based on a small study of 247 pregnant women with glucose >5.1 with only 4 cases of *GCK*-MODY, in contrast we had 163 cases of *GCK*-MODY with sample size of 454,275 potentially providing a more accurate prevalence estimate. The overall MODY prevalence in our population cohort was 1:1052 (0.095%), nearly three times higher than a previous estimate of ∼1:3521 (284 cases/million) in the UK (33). This difference likely reflects phenotype-based estimation of this study. We also found that MODY accounted for 1.48% before age 40, which is lower than the previously reported estimate of 3.6% in <30 years of age (34). This difference is potentially due to healthy participant bias in the UK Biobank, leading to likely lower representation of young-onset diseases, and the inclusion of m.3243A>G, and lipodystrophy genes in previous studies (35,36). A significantly lower prevalence of pathogenic MODY variants in individuals of African ancestry, has been reported with other monogenic diseases (37,38). This likely reflects the lack of sufficient available genetic evidence to classify novel or previously observed variants as pathogenic in this ancestry. Finally, the lack of female excess in our study, despite higher female participation in UK Biobank, strongly suggests that female overrepresentation in clinically ascertained cohorts results from an ascertainment bias.

Carriers of pathogenic *GCK* variants do not show excess all-cause mortality. Previous clinically ascertained cases demonstrated that *GCK*-MODY has high fasting glucose that increases with age (31). Here, using a large number of clinically unselected cases (163 individuals: mean age 56.8yrs), we observed similar findings. We also found that despite an average HbA1c of 47.1 mmol/mol, these individuals did not show excess all-cause mortality. These findings strongly support current guidelines which suggest discharging individuals with *GCK*-MODY from follow-up. Additionally, we did not observe excess mortality with any MODY subtype. This is particularly of note for *HNF1A*, where our findings contrast with previous studies. Steele *et al.* showed that clinically ascertained families with the *HNF1A* p.Pro291fsinsC variant had increased overall and cardiovascular mortality (12). Similar observation of all-cause mortality reported in Icelandic population in clinically unselected population for this particular variant (39). The discrepancy in results may be due to multiple factors. The healthy participant bias and minimum recruitment age of 40 years in the UK Biobank means young onset cases who died before 40 or severe cases would not be in the UK Biobank. The substantially improved cardiovascular preventative therapy and lower cardiovascular mortality over time in the UK general population may have lowered mortality in our cases (40,41), and finally, difficulties in accurately calling p.Pro291fsinsC variant from exome sequencing meant our study may have underrepresented this variant (only 16/60 individuals were confirmed to have the variant on manual check using IGV), which could have reduced the overall effect of *HNF1A* variants on all-cause mortality (10,11).

Our genotype-first approach shows that non-*GCK* MODY variants have variable penetrance and allowed us a unique insight into the heterogeneity of diabetes risk within the known MODY genes. We demonstrated that MODY variants increase diabetes risk as expected, but the absolute risk is much lower than clinically ascertained cases and varies substantially by genetic aetiology. We observed that *HNF1A* was the most penetrant with 60.6% developing diabetes by age 60yrs, whereas *ABCC8*, *HNF1B*, and *HNF4A* genes showed moderate penetrance (32.9-54.2% by 60yrs) and *NEUROD1*, *PDX1*, and *RFX6* genes showed the lowest penetrance (12-43.8% by 60yrs). This lower penetrance in clinically unselected cohorts has been observed previously for *HNF1A* and *HNF4A* (10,14). In this study, we extend this observation to all MODY genes. The lower penetrance in clinically unselected cohorts likely represents the lower bound of true penetrance, whereas clinical cohorts show the upper bound. The wide difference in penetrance between phenotype first and genotype first studies is not limited to diabetes but has been reported in other monogenic disorders (42–45). This wide range in penetrance suggests multiple modifiers are likely at play. In our limited sample size, we demonstrated that parental diabetes and polygenic background of type 2 diabetes independently affect penetrance likely due to overlapping pathophysiological pathway, an observation previously reported in clinically ascertained cases (46,47).The lack of impact of T1DGRS and BMI on MODY penetrance suggests that potential modifiers need to share a pathophysiological pathway with MODY genetic causes, which primarily involve beta cell dysfunction rather than autoimmune or adipocyte function. Further studies with larger numbers are needed to assess which pathways of type 2 diabetes genetic risk underlie this effect.

Our study has several limitations. We used a large population cohort in our study, but the UK Biobank has shown to have healthy participant bias partly due to a minimum recruitment age of 40 years (35). This has likely caused an underestimation of penetrance, prevalence and mortality. Despite using the largest cohort to date, we were still limited by the number of pathogenic carriers in some genes. Our cohort predominantly contains individuals of European ancestry, which may limit the generalisability of our findings to other populations.

In summary, we showed that 1 in 1052 individuals in the population have a pathogenic MODY variant and that MODY accounts for 1.48% of diabetes cases under 40yrs, with no impact on all-cause mortality and substantially lower penetrance than previously thought.

## Data Availability

UK Biobank dataset is available to researchers from https://biobank.ctsu.ox.ac.uk. The variants used in this study are available in the manuscript.

## Acknowledgements

This research has been conducted using the UK Biobank Resource. This work was conducted under UK Biobank project number 103356. The current work is funded by Diabetes UK (19/0005994 and 21/0006335), MRC (MR/T00200X/1). K.A.P is funded by the Wellcome Trust (219606/Z/19/Z). The work is supported by the National Institute for Health Research (NIHR) Exeter Biomedical Research Centre, Exeter, UK. The Wellcome Trust, MRC and NIHR had no role in the design and conduct of the study; collection, management, analysis, and interpretation of the data; preparation, review, or approval of the manuscript; and decision to submit the manuscript for publication. T.W.L is supported by the Academy of Medical Sciences/the Wellcome Trust/the Government Department of Science Innovation and Technology/the British Heart Foundation/Diabetes UK Springboard Award [SBF009\1135]. The views expressed are those of the author(s) and not necessarily those of the Wellcome Trust, Department of Health, NHS or NIHR. For the purpose of open access, the author has applied a CC BY public copyright licence to any Author Accepted Manuscript version arising from this submission. All authors declare no conflict of interest for this work.

