## Supplemental Tables and Figures for "Population prevalence, penetrance, and mortality for genetically confirmed MODY"

### Supplementary Materials

**Supplemental Table S1: Cohort characteristics of the UK Biobank**

| Characteristic | Value |
|---|---|
| N | 454,275 |
| Females, n (%) | 246,437 (54.25%) |
| Diabetes, n (%) | 28,795 (6.34%) |
| Age at recruitment (Years) | 57 (8.1) |
| BMI (kg/m$^2$) | 27.4 (4.8) |
| HbA1c (mmol/mol) | 38.4 (6.4) |
| Age of diabetes diagnosis (Years) | 55.9 (11.7) |
| Parent with diabetes, n (%) | 78,669 (17.32%) |
| European, n (%) | 419,796 (92.4%) |

Continuous traits are in mean and SD.

#### Supplemental Table S2: Pathogenic and Likely Pathogenic MODY variants from the UK Biobank.

| Genomic Change | Gene | Protein Change | Nucleotide Change | Criteria | Classification |
|---|---|---|---|---|---|
| 11:17460582:C:T | *ABCC8* | p.Arg306His | NM_001287174.1:c.917G>A | PM2 PP3 PS4 PP4_Moderate PM3 | Pathogenic |
| 11:17397788:C:T | *ABCC8* | p.Gly1256Ser | NM_001287174.1:c.3766G>A | PM2 PP3 PM6 PS4_Moderate PM3 PP4_Moderate | Likely Pathogenic |
| 11:17460529:C:T | *ABCC8* | p.Val324Met | NM_001287174.1:c.970G>A | PM2 PP3 PS4_Strong PP4_Moderate PM3 PS2 | Pathogenic |
| 11:17404525:G:A | *ABCC8* | p.Arg1183Trp | NM_001287174.1:c.3547C>T | PM2 PP3 PS4_Strong PP4_Moderate PM3 PM5 PP1_Strong PS2 | Pathogenic |
| 11:17393079:C:T | *ABCC8* | p.Arg1554Gln | NM_001287174.1:c.4661G>A | PM2 BP4 PP4_Moderate PS4_Moderate | Likely Pathogenic |
| 11:17397719:C:T | *ABCC8* | p.Gly1279Ser | NM_001287174.1:c.3835G>A | PM2 PS4_Supporting PP3 PP4_Moderate | Likely Pathogenic |
| 11:17413396:G:A | *ABCC8* | p.Arg826Trp | NM_001287174.1:c.2476C>T | PM2 PP3 PS4_Strong PP4_Moderate PM3 PP1_Strong | Pathogenic |
| 7:44151069:C:T | *GCK* | p.Asp124Asn | NM_000162.5:c.370G>A | PM2 PP3 PP1_Strong PS4_Moderate PM5_Supporting PP4 PP2 | Pathogenic |
| 7:44145270:G:A | *GCK* | p.Arg422Trp | NM_000162.5:c.1264C>T | PM2 PP3 PP4 PS4_Supporting PP1 PP2 PM5_Supporting | Likely Pathogenic |
| 7:44149977:G:A | *GCK* | p.Arg191Trp | NM_000162.5:c.571C>T | PS4 PS3 PP1_Strong PM6 PP4_Moderate PM2 PP2 PP3 | Pathogenic |
| 7:44147732:C:T | *GCK* | p.Gly261Arg | NM_000162.5:c.781G>A | PM2 PP2 PP3 PP4_Moderate PS3_Moderate PM3 PP1_Strong PS4 PS2 | Pathogenic |
| 7:44149763:C:T | *GCK* | p.Val226Met | NM_000162.5:c.676G>A | PM2_Supporting PP3 PS4 PM3_Supporting PP1_Strong | Pathogenic |
| 7:44146619:C:T | *GCK* | p.? | NM_000162.5:c.864-1G>A | PVS1 PM2 | Pathogenic |
| 7:44150004:C:T | *GCK* | p.Val182Met | NM_000162.5:c.544G>A | PM2 PP1_Strong PS4 PM5_Supporting PP4 PP2 PS3 | Pathogenic |
| 7:44149797:G:GT | *GCK* | p.Tyr214* | NM_000162.5:c.641dup | PVS1 PM2 PS4_Supporting PP1_Strong PP4 | Pathogenic |
| 7:44146531:G:C | *GCK* | p.His317Gln | NM_000162.5:c.951C>G | PM2 PP3 PP1_Moderate PS4_Moderate PM5_Supporting PP4 PP2 | Likely Pathogenic |
| 7:44149860:C:T | *GCK* | p.? | NM_000162.5:c.580-1G>A | PVS1 PM2 PP3 PS4_Supporting PP4 | Pathogenic |
| 7:44150978:A:G | *GCK* | p.Val154Ala | NM_000162.5:c.461T>C | PM2 PP3 PP1_Supporting PS4_Moderate PP4 PP2 | Likely Pathogenic |
| 7:44153326:G:T | *GCK* | p.Tyr61* | NM_000162.5:c.183C>A | PVS1 PM2 PP1_Strong PS4 PP4 | Pathogenic |
| 7:44145188:G:A | *GCK* | p.Ala449Val | NM_000162.5:c.1346C>T | PM2_Supporting PP2 PP3 PM5 PP4_Supporting | Likely Pathogenic |
| 7:44145522:C:G | *GCK* | p.Gly410Arg | NM_000162.5:c.1228G>C | PM2 PP3 PM5 PP4 | Likely Pathogenic |
| 7:44145176:G:A | *GCK* | p.Ser453Leu | NM_000162.5:c.1358C>T | PM2 PP3 PP1 PS4_Supporting PM5_Supporting PP4 PP2 PS3_Supporting | Likely Pathogenic |
| 7:44150008:A:C | *GCK* | p.Asn180Lys | NM_000162.5:c.540T>G | PM2 PP3 PS4 PP1_Strong | Pathogenic |
| 7:44150961:C:T | *GCK* | p.Asp160Asn | NM_000162.5:c.478G>A | PM2 PP1_Strong PS4 PM5_Supporting PP4 PP2 PS3 | Pathogenic |
| 7:44145212:G:A | *GCK* | p.Ser441Leu | NM_000162.5:c.1322C>T | PM2 PP3 PP2 PP4 PM5_Supporting | Likely Pathogenic |
| 7:44153372:C:A | *GCK* | p.Arg46Met | NM_000162.5:c.137G>T | PM2 PP3 PP2 PP4 PS4_Supporting | Pathogenic |
| 7:44149859:CCTGCCAAGAAGCA:C | *GCK* | p.? | NM_000162.5:c.580-13_580-1del | PVS1 PM2 PP1_Strong PP4 | Pathogenic |
| 7:44153381:C:T | *GCK* | p.Arg43His | NM_000162.5:c.128G>A | PS4_Moderate PP1_Moderate PP2 PP3 PM2 PM5 PP4 PS3 | Pathogenic |
| 7:44149992:G:A | *GCK* | p.Arg186* | NM_000162.5:c.556C>T | PVS1 PM2 PP1_Strong PS4 PP4 | Pathogenic |
| 7:44153299:A:G | *GCK* | p.? | NM_000162.5:c.208+2T>C | PVS1 PM2 PP3 | Pathogenic |
| 7:44149794:G:C | *GCK* | p.Tyr215* | NM_000162.5:c.645C>G | PVS1 PM2 PS4_Moderate PP1_Strong | Pathogenic |
| 7:44146604:A:G | *GCK* | p.Ile293Thr | NM_000162.5:c.878T>C | PM2 PP3 PM5_Supporting PS4_Moderate | Likely Pathogenic |
| 7:44146604:A:C | *GCK* | p.Ile293Arg | NM_000162.5:c.878T>G | PM2 PP3 PP1_Strong PS4_Moderate PM5 PP4 PP2 | Likely Pathogenic |
| 7:44145576:G:A | *GCK* | p.Arg392Cys | NM_000162.5:c.1174C>T | PP2 PP3 PP4 PM2 PS4 PP1_Strong | Pathogenic |
| 7:44152318:G:A | *GCK* | p.Gln106* | NM_000162.5:c.316C>T | PVS1 PM2 PP1 PS4_Moderate PP4 | Pathogenic |
| 7:44147765:G:A | *GCK* | p.Arg250Cys | NM_000162.5:c.748C>T | PS4_Supplemntary PP1_Strong PP4_Moderate PM2 PP2 PP3 | Pathogenic |
| 7:44145602:G:A | *GCK* | p.Ser383Leu | NM_000162.5:c.1148C>T | PM2 PP3 PS4 | Likely Pathogenic |
| 7:44147741:C:T | *GCK* | p.Gly258Ser | NM_000162.5:c.772G>A | PM2 PP3 PS4_Moderate PM5_Supporting | Likely Pathogenic |

| | | | | | |
|---|---|---|---|---|---|
| 7:44145608:A:G | *GCK* | p.Met381Thr | NM_000162.5:c.1142T>C | PM2 PP3 PS4 PM5 PP1 | Pathogenic |
| 7:44146580:A:G | *GCK* | p.Leu301Pro | NM_000162.5:c.902T>C | PM2 PP2 PP3 PP1_Strong PP4 | Likely Pathogenic |
| 7:44150961:C:A | *GCK* | p.Asp160Tyr | NM_000162.5:c.478G>T | PM2 PP3 PM5 PS4_Supporting | Likely Pathogenic |
| 7:44145228:T:A | *GCK* | p.Ile436Phe | NM_000162.5:c.1306A>T | PM2 PP3 PS4 | Likely Pathogenic |
| 7:44149838:C:A | *GCK* | p.Ala201Ser | NM_000162.5:c.601G>T | PM2 PP3 PS4_Moderate PP1_Strong | Likely Pathogenic |
| 7:44149986:C:T | *GCK* | p.Ala188Thr | NM_000162.5:c.562G>A | PM2 PP1_Strong PS4 PM5_Supporting PP4 PP2 PS3 | Pathogenic |
| 7:44152426:C:T | *GCK* | p.? | NM_000162.5:c.209-1G>A | PVS1 PM2 PS4_Supporting PP4 | Pathogenic |
| 7:44149794:G:T | *GCK* | p.Tyr215* | NM_000162.5:c.645C>A | PVS1 PM2 PS4 PP1_Strong | Pathogenic |
| 7:44145618:C:T | *GCK* | p.Ala378Thr | NM_000162.5:c.1132G>A | PM2 PP3 PS4_Moderate PM5 | Likely Pathogenic |
| 7:44149778:C:T | *GCK* | p.Glu221Lys | NM_000162.5:c.661G>A | PM2 PP1_Strong PS4 PP4_Moderate PP2 PS3 | Pathogenic |
| 7:44150954:A:G | *GCK* | p.? | NM_000162.5:c.483+2T>C | PVS1 PM2 PP3 PP4 PS4_Supporting | Pathogenic |
| 7:44147720:C:T | *GCK* | p.Glu265Lys | NM_000162.5:c.793G>A | PM2 PP3 PS4_Moderate PS3 | Pathogenic |
| 7:44149779:G:T | *GCK* | p.Cys220* | NM_000162.5:c.660C>A | PVS1 PM2 PS4_Moderate PP1_Strong | Pathogenic |
| 7:44153433:G:A | *GCK* | p.Gln26* | NM_000162.5:c.76C>T | PVS1 PM2 PP1 PS4 | Pathogenic |
| 7:44149813:G:A | *GCK* | p.Thr209Met | NM_000162.5:c.626C>T | PM2 PP3 PS4 | Likely Pathogenic |
| 7:44153379:C:T | *GCK* | p.Gly44Ser | NM_000162.5:c.130G>A | PM2 PP3 PP1_Strong PS4 PM5 PP4 PP2 | Pathogenic |
| 7:44149772:C:T | *GCK* | p.Gly223Ser | NM_000162.5:c.667G>A | PM2 PP3 PS4 PM3 PP1_Strong | Pathogenic |
| 7:44146463:C:A | *GCK* | p.Ser340Ile | NM_000162.5:c.1019G>T | PM2 PP3 PP1_Strong PS4_Moderate PM5 PP4 PP2 | Pathogenic |
| 7:44147678:C:A | *GCK* | p.Glu279* | NM_000162.5:c.835G>T | PVS1 PM2 PS4_Supporting PP1_Moderate | Pathogenic |
| 7:44152268:CA:C | *GCK* | p.? | NM_000162.5:c.363+2del | PVS1 PM2 | Pathogenic |
| 7:44146461:A:G | *GCK* | p.? | NM_000162.5:c.1019+2T>C | PVS1 PM2 PS3 PM3 PP1_Strong | Pathogenic |
| 7:44147690:G:C | *GCK* | p.Arg275Gly | NM_000162.5:c.823C>G | PM2 PP3 PM5 PP2 PP4_Supporting | Likely Pathogenic |
| 7:44150970:C:T | *GCK* | p.Glu157Lys | NM_000162.5:c.469G>A | PM2 PS4 | Likely Pathogenic |
| 7:44146619:C:G | *GCK* | p.? | NM_000162.5:c.864-1G>C | PVS1 PM2 PP3 | Pathogenic |
| 7:44151053:C:T | *GCK* | p.Cys129Tyr | NM_000162.5:c.386G>A | PM2 PP3 PS4 | Likely Pathogenic |
| 7:44153351:G:A | *GCK* | p.Ala53Val | NM_000162.5:c.158C>T | PM2 PP3 PP1_Strong PS4_Moderate PM5_Supporting PP4 PP2 | Pathogenic |
| 7:44147747:C:T | *GCK* | p.Glu256Lys | NM_000162.5:c.766G>A | PM2 PP3 PM5_Supporting PS4 PM3 | Pathogenic |
| 7:44151048:A:G | *GCK* | p.Ser131Pro | NM_000162.5:c.391T>C | PM2 PP1_Strong PP4 PP2 PS3_Supporting | Likely Pathogenic |
| 7:44149968:C:T | *GCK* | p.? | NM_000162.5:c.579+1G>A | PVS1 PM2 | Pathogenic |
| 7:44152275:TCAGCAGTG:T | *GCK* | p.Thr118Aspfs*8 | NM_000162.5:c.351_358del | PVS1 PM2 PP1 PS4_Moderate PP4 | Pathogenic |
| 7:44153325:C:T | *GCK* | p.Val62Met | NM_000162.5:c.184G>A | PM2 PP3 PP1_Strong PS3 PS4_Moderate | Likely Pathogenic |
| 7:44153379:C:A | *GCK* | p.Gly44Cys | NM_000162.5:c.130G>T | PM2 PP3 PM5 PP2 PP4 | Likely Pathogenic |
| 12:120997560:C:T | *HNF1A* | p.Gln466* | NM_000545.4:c.1396C>T | PVS1 PM2 PP1 PP4 PS3 | Pathogenic |
| 12:120994313:G:GC | *HNF1A* | p.Pro289Alafs*28 | NM_000545.4:c.863_864insC | PVS1 PS4_Moderate | Pathogenic |
| 12:120993584:G:T | *HNF1A* | p.Lys197Asn | NM_000545.4:c.591G>T | PM2 PS4_Supporting PP4_Moderate PP1_Supporting | Likely Pathogenic |
| 12:120988897:C:T | *HNF1A* | p.Arg131Trp | NM_000545.4:c.391C>T | PP3 PM1 PM2 PM5 PS4 PP1_Strong | Pathogenic |
| 12:120997639:C:T | *HNF1A* | p.Thr492Ile | NM_000545.4:c.1475C>T | PM2 PP3 PP1_Moderate PS4_Supporting | Likely Pathogenic |
| 12:120993678:C:T | *HNF1A* | p.Arg229* | NM_000545.4:c.685C>T | PVS1 PM2 PS4 PP1_Strong PP4 | Pathogenic |
| 12:120988981:C:T | *HNF1A* | p.Arg159Trp | NM_000545.4:c.475C>T | PP3 PP4 PM1_Supporting PM2 PM5 PS4 PP1_Strong | Pathogenic |
| 12:120993679:G:A | *HNF1A* | p.Arg229Gln | NM_000545.4:c.686G>A | PS3_Supporting PP1_Strong PS4 PM1_Supporting PM2 PP4_Moderate PP3 | Pathogenic |
| 12:120996569:C:G | *HNF1A* | p.Pro379Arg | NM_000545.4:c.1136C>G | PM2 PP3 PS3_Moderate PS4 | Likely Pathogenic |
| 12:120988849:G:T | *HNF1A* | p.Val115Leu | NM_000545.4:c.343G>T | PM2 PP3 PP4 PS4_Moderate PM5 | Likely Pathogenic |
| 12:120988909:GA:G | *HNF1A* | p.Asp135Valfs*20 | NM_000545.4:c.404del | PVS1 PM2 PS4 PP1_Strong | Pathogenic |

| | | | | | |
|---|---|---|---|---|---|
| 12:120989032:C:T | *HNF1A* | p.Gln176* | NM_000545.4:c.526C>T | PVS1 PM2 PS4 PP1_Supporting PP4_Moderate | Pathogenic |
| 12:120988853:C:T | *HNF1A* | p.Ala116Val | NM_000545.4:c.347C>T | PM2 PP3 PP1_Strong PS4 | Pathogenic |
| 12:120997491:GCA:G | *HNF1A* | p.Gln444Glufs*104 | NM_000545.4:c.1330_1331del | PVS1 PM2 PP4_Moderate PS4_Moderate PP1 | Pathogenic |
| 12:120994314:G:GC | *HNF1A* | p.Gly292Argfs*25 | NM_000545.4:c.872dup | PVS1 PS4 | Pathogenic |
| 12:120988898:G:A | *HNF1A* | p.Arg131Gln | NM_000545.4:c.392G>A | PM2 PP3 PM5 PS4_Moderate PS2 | Pathogenic |
| 12:120993601:G:A | *HNF1A* | p.Arg203His | NM_000545.4:c.608G>A | PM2_Supporting PP3 PS4 PP1_Moderate | Likely Pathogenic |
| 12:120993591:C:T | *HNF1A* | p.Arg200Trp | NM_000545.4:c.598C>T | PS4 PP1_Strong PM2 PM5 PP3 PP4 | Pathogenic |
| 12:120978928:C:T | *HNF1A* | p.Arg54* | NM_000545.4:c.160C>T | PVS1 PM2 PS4 PS2 PP1_Moderate | Pathogenic |
| 12:120996743:G:A | *HNF1A* | p.? | NM_000545.4:c.1309+1G>A | PVS1 PM2 | Pathogenic |
| 12:120993639:C:T | *HNF1A* | p.Gln216* | NM_000545.4:c.646C>T | PVS1 PM2 PS4_Supporting | Pathogenic |
| 12:120993592:G:A | *HNF1A* | p.Arg200Gln | NM_000545.4:c.599G>A | PM2 PP3 PM5 PS4 PP1_Strong | Pathogenic |
| 12:120994262:G:A | *HNF1A* | p.Arg271Gln | NM_000545.4:c.812G>A | PS4_Moderate PP1_Strong PM1 PP3 PM2 | Pathogenic |
| 12:120993519:G:A | *HNF1A* | p.? | NM_000545.4:c.527-1G>A | PVS1 PM2 PP3 PP4 PS4_Moderate PP1_Strong | Pathogenic |
| 12:120997647:CAGCTGCAG:C | *HNF1A* | p.Leu496Profs*50 | NM_000545.4:c.1487_1494del | PVS1 PM2 | Pathogenic |
| 12:120988937:T:C | *HNF1A* | p.Leu144Pro | NM_000545.4:c.431T>C | PM2 PP3 PS4 | Likely Pathogenic |
| 12:120994267:AAAG:A | *HNF1A* | p.Glu275del | NM_000545.4:c.824_826del | PM2 PM4_Supporting PS4 PP1_Strong | Pathogenic |
| 17:37731733:G:T | *HNF1B* | p.Arg303Ser | NM_000458.2:c.907C>A | PM2 PP3 PM5 PP4 | Likely Pathogenic |
| 17:37731599:C:CA | *HNF1B* | p.Ser348Valfs*12 | NM_000458.2:c.1040dup | PVS1 PM2 | Pathogenic |
| 17:37739491:G:A | *HNF1B* | p.Arg165Cys | NM_000458.2:c.493C>T | PM2 PP3 PM5 PP4 | Likely Pathogenic |
| 17:37739508:G:A | *HNF1B* | p.Pro159Leu | NM_000458.2:c.476C>T | PM2 PP3 PP1_Strong | Likely Pathogenic |
| 17:37687394:T:C | *HNF1B* | p.? | NM_000458.2:c.1654-2A>G | PVS1 PM2 | Pathogenic |
| 20:44413708:C:T | *HNF4A* | p.Arg112Trp | NM_175914.4:c.334C>T | PM2 PP3 PM5 PS4_Supplementary | Likely Pathogenic |
| 20:44413709:G:A | *HNF4A* | p.Arg112Gln | NM_175914.4:c.335G>A | PS4 PP1_Strong PP4_Moderate PM2 PP3 PM1 PM5_Supporting | Pathogenic |
| 20:44419837:G:C | *HNF4A* | p.Glu263Gln | NM_175914.4:c.787G>C | PP1_Strong PP4_Moderate PM2 PP3 PS3_Supporting | Likely Pathogenic |
| 20:44424147:T:C | *HNF4A* | p.Leu319Pro | NM_175914.4:c.956T>C | PM2 PP3 PS4_Supplementary PP1_Strong | Likely Pathogenic |
| 20:44424116:C:T | *HNF4A* | p.Arg309Cys | NM_175914.4:c.925C>T | PS4 PM1_Supporting PP1_Moderate PP4_Moderate PP3 | Pathogenic |
| 20:44413696:G:A | *HNF4A* | p.Val108Ile | NM_175914.4:c.322G>A | PM1_Supporting PM2 PP3 PS4_Moderate PP1_Strong PP4 | Pathogenic |
| 20:44424117:G:A | *HNF4A* | p.Arg309His | NM_175914.4:c.926G>A | PM1_Supporting PM2 PP3 PM5_Supporting PP4 PP1_Moderate | Likely Pathogenic |
| 20:44406132:G:A | *HNF4A* | p.Gly42Arg | NM_175914.4:c.124G>A | PP1 PP3 PM1_Supporting PM2 PS4_Supplementary | Likely Pathogenic |
| 20:44424117:G:T | *HNF4A* | p.Arg309Leu | NM_175914.4:c.926G>T | PM1_Supporting PM2 PP3 PM5_Supporting PP4_Moderate PP1_Moderate PS4_Moderate | Likely Pathogenic |
| 20:44406208:G:A | *HNF4A* | p.Arg67Gln | NM_175914.4:c.200G>A | PS4_Moderate PM2 PP3 PP1_Strong PP4_Moderate PM1 PM5_Supporting | Pathogenic |
| 20:44418456:A:C | *HNF4A* | p.His205Pro | NM_175914.4:c.614A>C | PP3 PM2 PP4_Moderate PP1 | Likely Pathogenic |
| 20:44355841:G:T | *HNF4A* | p.Glu13* | NM_175914.4:c.37G>T | PVS1 PM2 PP1 PS4_Supporting | Pathogenic |
| 20:44413726:C:T | *HNF4A* | p.Arg118* | NM_175914.4:c.352C>T | PVS1 PM2 PP4_Moderate PS4_Moderate PP1_Strong | Pathogenic |
| 20:44424224:G:T | *HNF4A* | p.Asp345Tyr | NM_175914.4:c.1033G>T | PP3 PM2 PS4_Supporting PP1 PP4 PS2 | Pathogenic |
| 20:44419783:C:T | *HNF4A* | p.Arg245Cys | NM_175914.4:c.733C>T | PM2 PP3 PM5_Supporting PM6 PP4_Supporting | Likely Pathogenic |
| 20:44419873:C:T | *HNF4A* | p.Pro275Ser | NM_175914.4:c.823C>T | PP3 PM2 PP4_Moderate PP1 | Likely Pathogenic |
| 20:44419741:C:T | *HNF4A* | p.Arg231Trp | NM_175914.4:c.691C>T | PP3 PM2 PM5 PS4_Moderate PP4 | Likely Pathogenic |
| 20:44424123:G:A | *HNF4A* | p.Arg311His | NM_175914.4:c.932G>A | PM2 PP3 PM5 PP4 PM1_Supporting | Likely Pathogenic |
| 20:44414610:T:C | *HNF4A* | p.Val177Ala | NM_175914.4:c.530T>C | PP3 PM2 PS4_Moderate PP4 PS4_Supporting PM5_Supporting | Likely Pathogenic |
| 20:44418467:G:A | *HNF4A* | p.Gly209Arg | NM_175914.4:c.625G>A | PP3 PM2 PP4_Moderate PP1 | Likely Pathogenic |
| 20:44424060:G:A | *HNF4A* | p.Arg290His | NM_175914.4:c.869G>A | PP3 PM2 PP1_Strong PP4_Moderate PS4 | Pathogenic |
| 20:44419784:G:A | *HNF4A* | p.Arg245His | NM_175914.4:c.734G>A | PM2 PP3 PS4_Supporting PP4 PS3_Supporting | Likely Pathogenic |

| | | | | | |
|---|---|---|---|---|---|
| 20:44414549:A:C | *HNF4A* | p.Lys157Gln | NM_175914.4:c.469A>C | PP3 PM2 PP4_Supporting PM1_Supporting PP1_Supporting | Likely Pathogenic |
| 20:44414617:G:A | *HNF4A* | p.Trp179* | NM_175914.4:c.537G>A | PVS1 PM2 PP4 | Pathogenic |
| 2:181678244:T:TG | *NEUROD1* | p.His206Profs*38 | NM_002500.4:c.616dup | PVS1 PM2 PP1 PS4 | Pathogenic |
| 2:181678432:CAG:C | *NEUROD1* | p.Leu143Alafs*55 | NM_002500.4:c.427_428del | PVS1 PM2 PM3_Supporting | Pathogenic |
| 2:181678072:ATCAG:A | *NEUROD1* | p.Thr262Ilefs*68 | NM_002500.4:c.785_788del | PVS1 PM2 | Pathogenic |
| 2:181678168:G:T | *NEUROD1* | p.Tyr231* | NM_002500.4:c.693C>A | PVS1 PM2 | Pathogenic |
| 13:27920320:GC:G | *PDX1* | p.Pro63Argfs*60 | NM_000209.3:c.188del | PVS1 PM2 PS4_Supporting | Pathogenic |
| 13:27920208:C:CCGGCGCCGGAGTT | *PDX1* | p.Ser29Glyfs*200 | NM_000209.3:c.72_84dup | PVS1 PM2 | Pathogenic |
| 13:27920482:T:TC | *PDX1* | p.Gln116Profs*109 | NM_000209.3:c.346dup | PVS1 PM2 | Pathogenic |
| 13:27920410:C:CG | *PDX1* | p.Leu92Alafs*133 | NM_000209.3:c.273dup | PVS1 PM2 | Pathogenic |
| 13:27920188:C:CA | *PDX1* | p.Cys18Metfs*207 | NM_000209.3:c.51dup | PVS1 PM2 | Pathogenic |
| 13:27920230:GC:G | *PDX1* | p.Pro33Leufs*90 | NM_000209.3:c.98del | PVS1 PM2 | Pathogenic |
| 13:27920342:C:A | *PDX1* | p.Tyr68* | NM_000209.3:c.204C>A | PVS1 PM2 | Pathogenic |
| 13:27920443:AG:A | *PDX1* | p.Gly103Glufs*20 | NM_000209.3:c.308del | PVS1 PM2 | Pathogenic |
| 6:116882403:C:T | *RFX6* | p.Arg181Trp | NM_173560.3:c.541C>T | PM2 PP3 PS4_Moderate PP4 PM3_Supporting | Likely Pathogenic |
| 6:116916208:A:AT | *RFX6* | p.Leu291Phefs*10 | NM_173560.3:c.872dup | PVS1 PM2 | Pathogenic |
| 6:116920455:G:A | *RFX6* | p.? | NM_173560.3:c.1327+1G>A | PVS1 PM2 | Pathogenic |
| 6:116919165:A:T | *RFX6* | p.Lys351* | NM_173560.3:c.1051A>T | PVS1 PM2 | Pathogenic |
| 6:116927095:C:T | *RFX6* | p.Arg652* | NM_173560.3:c.1954C>T | PVS1 PM2_Supporting | Pathogenic |
| 6:116916217:T:G | *RFX6* | p.Leu292* | NM_173560.3:c.875T>G | PVS1 PM2 PP1_Moderate PS4_Supporting | Pathogenic |
| 6:116923154:TC:T | *RFX6* | p.Leu496Cysfs*2 | NM_173560.3:c.1486del | PVS1 PM2 | Pathogenic |
| 6:116916031:C:A | *RFX6* | p.Tyr268* | NM_173560.3:c.804C>A | PVS1 PM2 | Pathogenic |
| 6:116925497:CT:C | *RFX6* | p.Leu575Argfs*15 | NM_173560.3:c.1724del | PVS1 PM2 | Pathogenic |
| 6:116924686:C:T | *RFX6* | p.Arg525* | NM_173560.3:c.1573C>T | PVS1 PM2 PS4_Supporting PM3_Supporting | Pathogenic |
| 6:116877884:GTC:G | *RFX6* | p.Gln106Glufs*11 | NM_173560.3:c.315_316del | PVS1 PM2 | Pathogenic |
| 6:116877348:C:T | *RFX6* | p.Gln25* | NM_173560.3:c.73C>T | PVS1 PM2 PS4_Moderate | Pathogenic |
| 6:116919142:T:G | *RFX6* | p.Leu343* | NM_173560.3:c.1028T>G | PVS1 PM2 PS4_Supporting | Pathogenic |
| 6:116927106:C:CATTAACCA | *RFX6* | p.Gly659Leufs*117 | NM_173560.3:c.1967_1974dup | PVS1 PM2 | Pathogenic |
| 6:116928853:G:GC | *RFX6* | p.Tyr834Leufs*3 | NM_173560.3:c.2499dup | PVS1 PM2 PS4 | Pathogenic |
| 6:116919243:C:T | *RFX6* | p.Arg377* | NM_173560.3:c.1129C>T | PVS1 PM2 PS4_Supporting PM3 | Pathogenic |
| 6:116919215:TCTAA:T | *RFX6* | p.Asp370Argfs*13 | NM_173560.3:c.1104_1107del | PVS1 PM2 | Pathogenic |
| 6:116916251:T:TG | *RFX6* | p.Pro304Alafs*10 | NM_173560.3:c.909_910insG | PVS1 PM2 | Pathogenic |
| 6:116919157:AT:A | *RFX6* | p.Phe349Leufs*20 | NM_173560.3:c.1047del | PVS1 PM2 | Pathogenic |
| 6:116880544:G:A | *RFX6* | p.Trp127* | NM_173560.3:c.381G>A | PVS1 PM2 | Pathogenic |
| 6:116927431:C:T | *RFX6* | p.Gln764* | NM_173560.3:c.2290C>T | PVS1 PM2 | Pathogenic |
| 6:116916031:CA:C | *RFX6* | p.Thr270Leufs*74 | NM_173560.3:c.808del | PVS1 PM2 | Pathogenic |
| 6:116925596:G:T | *RFX6* | p.Gly608* | NM_173560.3:c.1822G>T | PVS1 PM2 | Pathogenic |
| 6:116877366:C:T | *RFX6* | p.Gln31* | NM_173560.3:c.91C>T | PVS1 PM2 | Pathogenic |
| 6:116919224:CAAGAA:C | *RFX6* | p.Lys371Asnfs*30 | NM_173560.3:c.1113_1117del | PVS1 PM2 | Pathogenic |
| 6:116927229:TTA:T | *RFX6* | p.Tyr697* | NM_173560.3:c.2091_2092del | PVS1 PM2 | Pathogenic |
| 6:116877450:GGC:G | *RFX6* | p.Lys62Argfs*20 | NM_173560.3:c.177_178del | PVS1 PM2 | Pathogenic |
| 6:116916214:T:A | *RFX6* | p.Leu291* | NM_173560.3:c.872T>A | PVS1 PM2 | Pathogenic |
| 6:116925452:G:A | *RFX6* | p.? | NM_173560.3:c.1679-1G>A | PVS1 PM2 | Pathogenic |
| 6:116919267:C:T | *RFX6* | p.Arg385* | NM_173560.3:c.1153C>T | PVS1 PM2 | Pathogenic |

| | | | | | |
|---|---|---|---|---|---|
| 6:116916014:AC:A | *RFX6* | p.Thr263Serfs*3 | NM_173560.3:c.788del | PVS1 PM2 | Pathogenic |
| 6:116894011:CAA:C | *RFX6* | p.Lys198Argfs*26 | NM_173560.3:c.593_594del | PVS1 PM2 | Pathogenic |
| 6:116927041:TCA:T | *RFX6* | p.Gln635Aspfs*22 | NM_173560.3:c.1903_1904del | PVS1 PM2 | Pathogenic |
| 6:116877864:A:T | *RFX6* | p.Lys98* | NM_173560.3:c.292A>T | PVS1 PM2 | Pathogenic |
| 6:116882419:GC:G | *RFX6* | p.His187Ilefs*27 | NM_173560.3:c.559del | PVS1 PM2 | Pathogenic |
| 6:116927491:G:GGAGCTGC | *RFX6* | p.Gly790Leufs*9 | NM_173560.3:c.2351_2357dup | PVS1 PM2 | Pathogenic |
| 6:116882404:G:A | *RFX6* | p.Arg181Gln | NM_173560.3:c.542G>A | PM2 PP3 PM5_Supporting PS3 | Likely Pathogenic |
| 6:116877318:C:T | *RFX6* | p.Gln15* | NM_173560.3:c.43C>T | PVS1 PM2 | Pathogenic |
| 6:116925508:T:TA | *RFX6* | p.Asn579Lysfs*2 | NM_173560.3:c.1736dup | PVS1 PM2 | Pathogenic |
| 6:116927460:TAC:T | *RFX6* | p.Thr774Argfs*22 | NM_173560.3:c.2321_2322del | PVS1 PM2 | Pathogenic |
| 6:116923168:G:GT | *RFX6* | p.Gly503Trpfs*7 | NM_173560.3:c.1506dup | PVS1 PM2 | Pathogenic |
| 6:116920308:A:C | *RFX6* | p.? | NM_173560.3:c.1183-2A>C | PVS1 PM2 | Pathogenic |
| 6:116882420:CCATT:C | *RFX6* | p.His187Glnfs*26 | NM_173560.3:c.561_564del | PVS1 PM2 | Pathogenic |
| 6:116877852:GAC:G | *RFX6* | p.Asp94Glufs*23 | NM_173560.3:c.282_283del | PVS1 PM2 | Pathogenic |

Variants were identified from the 450k exome release from the UKBB and was classified using the criteria from *Ellard et al.* 2020.

**Supplemental Table S3: Prevalence of pathogenic MODY variants by genetic ancestry.**

| Genetic Ancestry | Prevalence % (95%CI) | MODY N | Total Ancestry N | MODY N per 100,000 |
|---|---|---|---|---|
| European | 0.096 (0.087-0.106) | 404 | 419,796 | 92.4 |
| South Asian | 0.12 (0.058-0.21) | 11 | 9,537 | 115.3 |
| African* | 0.014 ($3.4x10^{-4}$-0.075) | 1 | 7,393 | 13.5 |
| Other | 0.091 (0.052-0.15) | 16 | 17,549 | 91.2 |

*Heterogeneity across ancestries $p=2.07x10^{-7}$. Heterogeneity across ancestry without Africans p =0.84

**Supplemental Table S4: Change in HbA1c and Random blood glucose between *GCK* Carriers and non-carriers with and without diabetes**

| | | ***GCK* vs All Non-Carriers** | | ***GCK* vs Non-Carriers without diabetes** | |
|---|---|---|---|---|---|
| **Blood Measurement Method** | **Adjusted/Unadjusted** | **Mean difference (95% CI)** | **P-value** | **Mean Difference (95% CI)** | **P-value** |
| HbA1c (mmol/mol) | Unadjusted* | 8.78(7.79-9.75) | $7.61x10^{-69}$ | 9.62(8.96-10.28) | $7.95x10^{-179}$ |
| | Adjusted† | 8.41(7.49-9.34) | $3.98x10^{-71}$ | 9.42(8.79-10.04) | $1.47x10^{-190}$ |
| Blood Glucose after 5hr fasting (mmol/L) | Unadjusted* | 1.28(0.96-1.61) | $5.4x10^{-15}$ | 1.38(1.17-1.59) | $1.87x10^{-38}$ |
| | Adjusted‡ | 1.28(0.97-1.6) | $1.56x10^{-15}$ | 1.38(1.17-1.58) | $2.08x10^{-39}$ |

Blood glucose was only used for individuals who had been fasting for > 5 hours. *Adjusted for genetic ancestry principal components. † Adjusted for age, parent diabetes status, BMI, sex, genetic ancestry principal components. ‡Adjusted for age, parent diabetes status, BMI, sex, genetic ancestry principal components and fasting time.

**Supplemental Table S5: Hazard ratio of diabetes for all MODY and by each genetic aetiology from a Cox proportional hazard model**

| Group | Unadjusted* | | | | Adjusted† | | | |
|---|---|---|---|---|---|---|---|---|
| | MODY N | Non-Carrier N | Hazard Ratio | p-value | MODY N | Non-Carrier N | Hazard Ratio | p-value |
| All Non-*GCK* MODY | 266 | 452760 | 6.86(5.55-8.48) | $4.19x10^{-71}$ | 264 | 450940 | 6.93(5.6-8.58) | $5.81x10^{-71}$ |
| *ABCC8* | 12 | 453170 | 20.82(9.35-46.36) | $1.05x10^{-13}$ | 12 | 451348 | 15.68(7.04-34.9) | $1.59x10^{-11}$ |
| *HNF1A* | 50 | 453132 | 26.01(18.39-36.79) | $9.71x10^{-76}$ | 50 | 451310 | 22.09(15.62-31.26) | $1.80x10^{-68}$ |
| *HNF1B* | 16 | 453166 | 11.97(5.7-25.1) | $5.17x10^{-11}$ | 16 | 451344 | 23.02(10.97-48.3) | $1.08x10^{-16}$ |
| *HNF4A* | 54 | 453128 | 6.53(4.21-10.13) | $4.99x10^{-17}$ | 53 | 451307 | 6.82(4.35-10.69) | $6.41x10^{-17}$ |
| *NEUROD1* | 26 | 453156 | 4.39(1.83-10.55) | $9.42x10^{-04}$ | 26 | 451334 | 3.28(1.36-7.88) | $7.92x10^{-03}$ |
| *PDX1* | 13 | 453169 | 5.86(2.2-15.62) | $4.07x10^{-04}$ | 13 | 451347 | 5.2(1.95-13.86) | $9.82x10^{-04}$ |
| *RFX6* | 95 | 453087 | 2.14(1.21-3.77) | $8.50x10^{-03}$ | 94 | 451266 | 2.34(1.33-4.12) | $3.21x10^{-03}$ |

*Adjusted only for genetic ancestry principal components. † Adjusted for Sex, Parents' diabetes status, age at recruitment, BMI and genetic ancestry principal components.

**Supplemental Table S6: Phenotypic and genetic factors affecting penetrance by MODY subtype groups**

| Factor | Low Penetrance Group Hazard Ratio (95%CIs) | Low Penetrance Group P-value | Moderate Penetrance Group Hazard Ratio (95%CIs) | Moderate Penetrance Group P-value | High Penetrance Group Hazard Ratio (95%CIs) | High Penetrance Group P-value |
|---|---|---|---|---|---|---|
| Parent with diabetes | 1.28(0.41-4.06) | 0.67 | 4.88(1.68-14.22) | $3.61x10^{-3}$ | 2.28(0.79-6.56) | 0.058 |
| T2DPRS* | 1.83(0.95-3.54) | 0.70 | 1.87(1.01-3.46) | 0.05 | 1.03(0.56-1.90) | 0.91 |

These multivariable cox proportional hazard models were adjusted for genetic ancestry principal components, genetic aetiology, age at recruitment, sex, BMI, parental diabetes status, T1DGRS and T2DGRS.The low penetrance group contains 129 carriers of pathogenic variants in *RFX6*, *NEUROD1* or *PDX1*. The moderate penetrance group contains 75 carriers of pathogenic variants in *HNF1B*, *HNF4A* and *ABCC8*. The high penetrance group contains 49 carriers of pathogenic variants in *HNF1A*. * T2DPRS is standardised.

**Supplemental Table S7: Adjusted all-cause mortality by genetic aetiology.**

| Group | Hazard ratio | p-value |
|---|---|---|
| Non-*GCK* MODY | 0.81(0.53-1.23) | 0.32 |
| *GCK* | 0.94(0.60-1.48) | 0.79 |
| *HNF1A* | 0.32(0.08-1.48) | 0.11 |
| *HNF1B* | 2.01(0.64-6.23) | 0.23 |
| *HNF4A* | 1.18(0.56-2.48) | 0.66 |
| *NEUROD1* | 0.75(0.19-3.00) | 0.69 |
| *PDX1* | 3.44(0.86-13.76) | 0.08 |
| *RFX6* | 0.66(0.30-1.48) | 0.32 |

Hazard ratios for all-cause mortality form a Cox proportional hazard model adjusted for sex, smoking status, diabetes status, age at recruitment ,HbA1c, genetic ancestry principal components. This analysis has all the non-*GCK* MODY genes combined in Non-*GCK* MODY and each subtype has been also analysed individually, except *ABCC8* which has been excluded due to no mortality in the carriers.

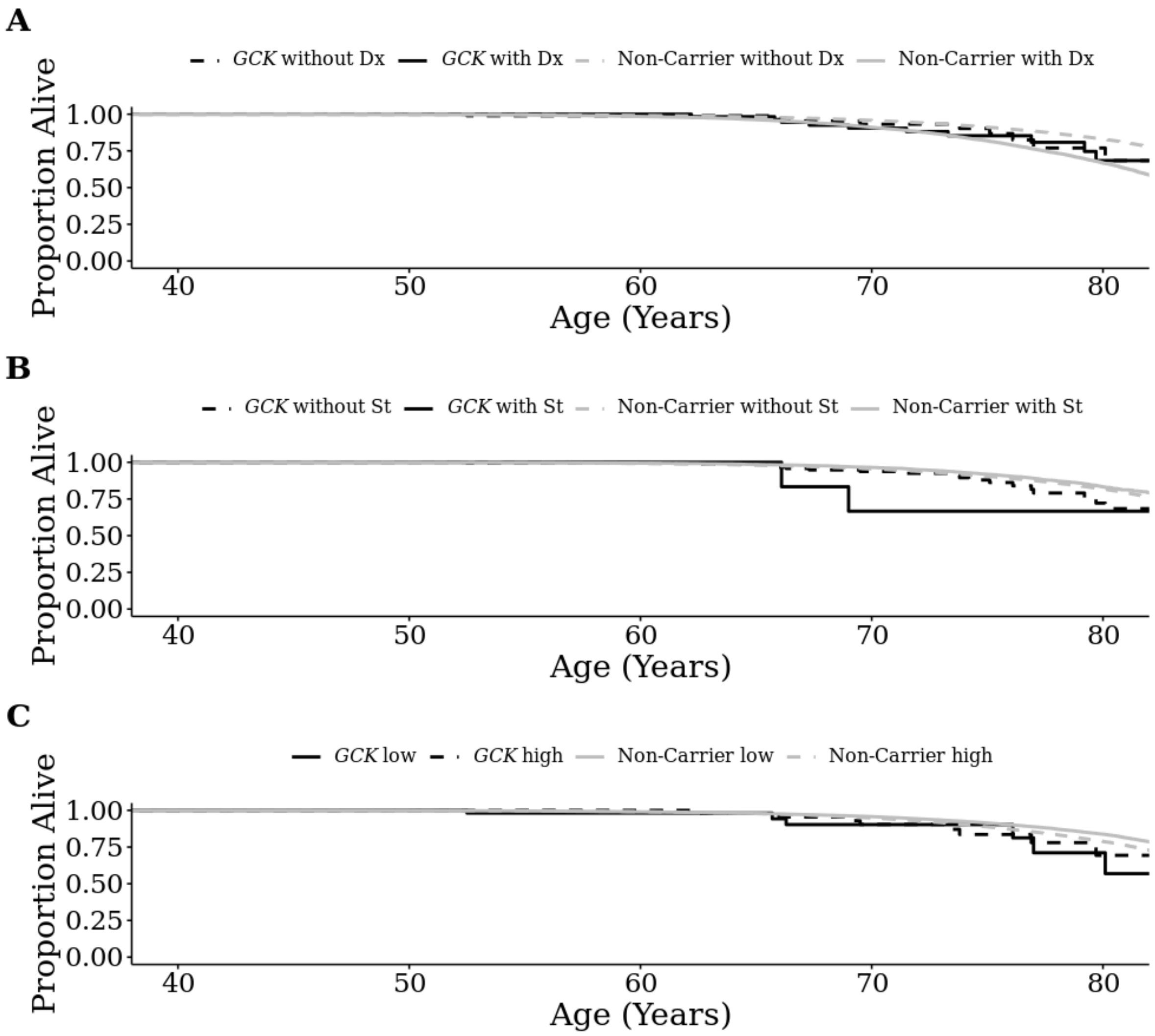


**Supplemental Figure S1: Kaplan Meier Curve for Mortality in *GCK*-MODY patients and individuals split my diabetes diagnosis by doctor, statin treatment status and high and low HbA1c tertile.** Kaplan-Meier analysis of Mortality stratified by *GCK* pathogenic variant carrier status and A) diabetes diagnosis (dx) status (*GCK* with dx = 64, *GCK* without dx = 92, Non-Carrier with dx = 23,647, Non-Carrier without dx = 428,499), B) statin (St) treatment status (*GCK* with statins = 8, *GCK* without statins = 155, Non-Carrier with statins = 8,199, Non-Carrier without statins = 445,913) and C) upper or lower HbA1c tertile (*GCK* lower HbA1c tertile = 54, *GCK* upper HbA1c tertile = 53, Non-Carrier lower HbA1c tertile = 144,036, Non-Carrier upper HbA1c tertile = 144,035 ).
